# Cross-continental environmental and genome-wide association study on children and adolescent anxiety and depression

**DOI:** 10.1101/2023.02.06.23285530

**Authors:** Bishal Thapaliya, Bhaskar Ray, Britny Farahdel, Pranav Suresh, Ram Sapkota, IMAGEN consortium, cVEDA consortium, Bharath Holla, Jayant Mahadevan, Jiayu Chen, Nilakshi Vaidya, Nora Perrone-Bizzozero, Vivek Benegal, Gunter Schumann, Vince D. Calhoun, Jingyu Liu

## Abstract

Anxiety and depression in children and adolescents warrant special attention as a public health issue given their devastating and long-term effects on development and mental health. Multiple factors, ranging from genetic vulnerabilities to environmental stressors, influence the risk for the disorders. This study aimed to understand how environmental factors and genomics affect children and adolescents anxiety and depression across three cohorts: Adolescent Brain and Cognitive Development Study (US, age of 9-10), Consortium on Vulnerability to Externalizing Disorders and Addictions (INDIA, age of 6-17) and IMAGEN (EUROPE, age of 14). We performed data harmonization and identified the environmental impact on anxiety/depression using a linear mixed-effect model, recursive feature elimination regression, and the LASSO regression model. Subsequently, genome-wide association analyses with consideration of significant environmental factors were performed for all three cohorts by mega-analysis and meta-analysis, followed by functional annotations. The results showed that multiple environmental factors contributed to the risk of anxiety and depression during development, where early life stress and school risk had the most significant and consistent impact across all three cohorts. Both meta and mega-analysis identified a novel SNP rs79878474 in chr11p15 to be the most promising SNP associated with anxiety and depression. Gene set analysis on the common genes mapped from top promising SNPs of both meta and mega analyses found significant enrichment in regions of chr11p15 and chr3q26, in the function of potassium channels and insulin secretion, in particular Kv3, Kir-6.2, SUR potassium channels encoded by the KCNC1, KCNJ11, and ABCCC8 genes respectively, in chr11p15. Tissue enrichment analysis showed significant enrichment in the small intestine and a trend of enrichment in the cerebellum. Our findings provide evidence of consistent environmental impact from early life stress and school risks on anxiety and depression during development and also highlight the genetic association between mutations in potassium channels along with the potential role of the cerebellum region, which are worthy of further investigation.

## Introduction

Anxiety and depression are now considered to be two of the most frequent mental disorders that affect children and adolescents^1^. The occurrence of anxiety and depression in children and adolescents, as well as other related mental disorders, is currently a worldwide pressing problem. According to the United States Centers of Disease Control National Survey of Children’s Health, 7.1% of children aged 3-17 years (about 4.4 million) have been diagnosed with anxiety, 3.2% have been diagnosed with depression (roughly 1.9 million)^2^, and this percentage increased to 11.7% for adolescents. The WHO has reported that one in every four children in India aged 13 to 15 suffers from depression. United Nations International Children’s Emergency Fund has reported that nine million adolescents in Europe (aged 10 to 19) are living with mental disorders, with anxiety and depression accounting for more than half of all cases (https://www.unicef.org/eu/stories/mental-health-burden-affecting-europes-children). In addition, a vast body of research from epidemiological surveys has shown a strong link between depression and anxiety with other mental disorders, particularly substance use disorders^3,4^. According to a major US survey, 14% of respondents with major depression reported an alcohol use problem in the previous 12 months, and 4.6% had a drug use disorder^5^. A Norwegian study also found that higher levels of depression symptoms were associated with earlier onset of alcohol use, more frequent consumption and intoxication^6^.

The causal pathways of anxiety and depression are not fully delineated yet, but the risk factors are multifaceted as shown in previous studies. Poverty^7,8^, dysfunctional family relationships and parental divorce^9,10^, child abuse^11,12^, and other stressful life events^13,14^ are well-known environmental risk factors for anxiety and depression. Furthermore, it has been discovered that teenagers who live in an area surrounded by trees and other green vegetation (i.e., green space) had a lower risk of severe depressive symptoms^15^. The impact of various levels of environmental factors from the individual micro level, to neighborhood middle and regional macro levels collectively in a broader setting across continents, has yet to be investigated to test the generalizability and specificity of environmental effects.

The largest genome-wide association study (GWAS) ever conducted for anxiety found substantial connections between self-reported anxiety and specific single nucleotide polymorphisms (SNPs) in a total of 200,000 participants^16^. Most of the identified risk SNPs are situated in non-coding areas, implying that these genetic variants may transmit the risk of anxiety disorders or traits by regulating gene expression^17–20^. Depression has a genetic component as well, with heritability estimated 31% to 42% in twin studies of children and adolescents^21^. Several large GWAS on depression have been recently conducted providing top-risk SNPs in general^22,23^. Additionally, substantial genetic correlations were observed between panic disorder and MDD, depressive symptoms, and neuroticism in a recent GWAS meta-analysis in European countries (Denmark, Estonia, Germany, and Sweden)^24^. However, there has not been a huge success in explicitly identifying the sensitive genes or genetic risks on adolescent depression and anxiety^25^, likely due to complicated genetic-environmental-developmental interactions. The current study is focused on understanding the genetic and environmental influence on anxiety and depression during development on a large geographic scale, with the hope to more clearly delineate the consistent, as well as unique genes and environmental effects across continents.

We have recently published a study^26^ using Adolescent Brain and Cognitive Development Study (ABCD) data to identify environmental and genetic risk factors for anxiety and depression in children. One overall score to represent combined anxiety and depression severity was chosen due to the highly common occurrence: about 3 in 4 children with depression also had anxiety^27^. The findings support that environmental factors from the personal level (early life stress, household income), to neighborhood level (school risk, area crime), and to the large scale of population density, all contribute to anxiety and depression in children. Together they could explain 6.2% of severity variance. Genetic variants also contribute to anxiety and depression, which could explain 10-15% of the severity variance measured by SNP heritability. With global mental health being a tremendous issue, we aim to study the effect of genetic and environmental factors across the US, India, and Europe and explore the general and specific effects. To our best knowledge, this is the first study that considered different levels of environmental factors when performing the GWAS of anxiety/depression in children and adolescents across three cohorts of very diverse backgrounds. Specifically, in the current study we characterize the impact of environmental factors on children and adolescents anxiety and depression and then perform GWAS to examine the influence of genetics with proper consideration of environmental factors. Both mega-analysis and meta-analysis are performed to integrate results from three cohorts, and followed by functional annotations for resultant SNPs, genes, and gene sets.

## Methods

### Data and Participants

In this study, we analyzed data from three big cohorts: ABCD from US, IMAGEN from Europe, and the Consortium on Vulnerability to Externalizing Disorders and Addictions (c-VEDA) from India. Participants from each cohort all signed the consent form for the original studies, and the original studies were approved by local ethic committees.

#### The ABCD Dataset

ABCD is one of the largest ongoing studies following youths recruited at age 9-10 into late adolescence^28^to broaden our understanding of emotional, genetic, neurological, and behavioral factors that are responsible to increase the risk of physical and mental health problems in youth. It is designed to run for at least 10 years and recruit participants from 21 sites across the United States. The recruitment catchment areas are believed to encompass over 20% of the entire 9-10-year-old population in the US on several key demographic variables, including gender, race/ethnicity, household income, parental education, and marital status. Further information on recruitment sites, study design, investigators, and partners can be obtained at http://abcdstudy.org. We used the data from ABCD Data Release 3.0, which is available on the NIMH Data Archive (https://nda.nih.gov/abcd). Assessments we analyzed include Parent-rated Child Behavior Checklist (CBCL), School Risk and Protective Factors Survey, Youth Family Environment Scale-Family Conflict, Longitudinal Parent Demographics Survey, Parent Neighborhood Safety/Crime Survey, Sum Scores Culture & Environment Youth, Residential History Derived Scores, and Youth Neighborhood Safety/Crime Survey. From a total of 11,875 samples at baseline (ages 9-10 years old), we removed samples with any missing values, resulting in 8,513 samples for further analyses.

#### The cVEDA Dataset

The c-VEDA is a cooperative initiative by the Medical Research Council, UK (MRC) and the Indian Council for Medical Research (ICMR) on the etiology and life-course of substance addiction and its link with mental illness (ICMR)^29^. The coordinating centers in India and the United Kingdom are the National Institute of Mental Health and Neurosciences in Bangalore (NIMHANS) and King’s College London (KCL), respectively. cVEDA has recruited participants with specific age ranges of 6-11, 12-17 and 18-23 years from seven centers of five geographical regions of India: Punjab and adjoining states (PGIMER), Eastern Coalfields (KOLKATA), Northeast India (IMPHAL), Bengaluru and Mysuru (MYSORE, NIMHANS, SJRI) and Chittoor (RISHIVALLEY). We analyzed data from the Mini-International Neuropsychiatric Interview - KID (MINI-KID), Environmental Exposures Questionnaire, Adverse Childhood Experiences International Questionnaire, Indian Family Violence, and Control Scale Questionnaire, Socioeconomic Status Questionnaire, and the School Experience Questionnaire to characterize environmental factors and anxiety/depression rate. Further information about the questionnaires can be found in the Supplementary files. We studied the data involving children (aged 6-11) and adolescents (aged 12-17). After removing the missing values, we had data from 4,326 samples.

#### The IMAGEN Dataset

The IMAGEN database contains data collected and processed by the IMAGEN consortium from over 2000 adolescents and their parents^30^. It includes demographics, neuropsychological assessments, medical questionnaires, MR neuroimaging and genomics. Data have been collected over a period of 10 years in eight recruitment centers and over four successive time points: baseline at age 14, follow-up 1 at age 16, follow-up 2 at age 19, and follow-up 3 at age 23. Life Events Questionnaire, Bully Questionnaire, and the Development and Well-Being Assessment Interview Questionnaire (DAWBA) were used from the IMAGEN cohort to identify the effect of anxiety/depression along with all other environmental factors. Further information about the questionnaires can be found in the Supplementary files. We used the baseline data at age 14, and with preprocessing by removing the missing values, the total number of samples was 1,888.

### Defining environmental Factors

Based on the availability of variables, we extracted environmental factors for all three datasets (ABCD, cVEDA and IMAGEN). We have used eight environmental factors (air pollution, population density, area crime, neighborhood safety, school risk, household income, family conflict, early life stress (ELS)) for the ABCD Cohort, five factors (air pollution, school risk, household income, family conflicts, ELS) for the cVEDA, and three factors (ELS, school risk, family conflicts) for the IMAGEN cohort. Each factor is derived from multiple variables assessing related issues. Specifically, related variables were summed together to get a more general measure for that particular environmental factor. All the details of the variables used and questions for each variable can be found in the Supplementary files.

### Defining the anxiety/depression score

In the ABCD study, the parent-rated CBCL is used to determine the rate of depression/anxiety in children. CBCL is a component of the Achenbach System of Empirically Based Assessment, which is designed to detect emotional and behavioral problems in children and adolescents. The behaviors of the child across the past six months were reported by the parent through 113 questions. We selected 13 variables from the CBCL to capture aspects of anxiety and depression. In cVEDA study, we used five variables in MINI-KID to identify the rate of Anxiety/Depression. In IMAGEN cohort, 62 variables in DAWBA were used to measure anxiety/depression scores. Finally, for all three cohorts, the sum of these variables was used to measure the overall score of anxiety and depression. See the exact questions used in the Supplementary file.

### Genomic data preprocessing

Genomic data were quality controlled to prevent spurious association detection. As ABCD provided imputed whole genome data in release 3.0, we used the data provided by the consortium where imputation was performed using the TOPMed imputation server following the pre-imputation steps as instructed at (https://topmedimpute.readthedocs.io/en/latest/prepare-your-data/). With same steps we performed the imputation for IMAGEN genomic data using the TOPMed Imputation Server^31^. Imputation of cVEDA genomic data was using the Michigan imputation server^32^ and the South Asian Ancestry (SAS) reference panel. LiftOver was performed to represent SNPs in HG38 coordinates using LiftOver in UCSC Genome Browser^33^. The results of imputation from both cVEDA and IMAGEN were thresholded with imputation R2>0.3. After imputation, further filtering steps were applied to SNPs including genotyping rate (missing rate per SNP) of 0.05, a minor allele frequency of 0.01, and a Hardy-Weinberg equilibrium threshold of 1e-06. Furthermore, the individuals with more than 3 standard deviations away from the samples’ heterozygosity rate mean were removed. Finally, we had 10908 subjects and 8812066 SNPs for ABCD, 1014 subjects and 4475075 SNPs for cVEDA, and 1831 subjects and 8785037 SNPs for IMAGEN respectively.

### Data Analyses

#### Data Harmonization with reference to ABCD Cohort

The current study intends to assess the general effect of each environmental factor on children’s anxiety and depression across the three cohorts. Achieving the goal requires data harmonization, as the utilized environmental and anxiety/depression assessments varied across the three cohorts. Data harmonization can generate comparable datasets from heterogeneous sources. Specifically, we compared the cumulative distribution function (CDF) of each factor. The CDF of random variable X is defined as *F*_*X*_ (*x*) = *P*(*X* ≤ *x*), for all *x* ∈ *R*, where *P*(*X* ≤ *x*) represents the probability that the random variable X takes on a value less than or equal to x. After scaling each factor into 0-1 range, we applied gamma transformation on cVEDA and IMAGEN factors using ABCD factors as references. Gamma transformation (power transformation) defined as *y* = *X*^*γ*^ is a monotonic transformation where *γ* is chosen so that the values of CDF at 90% of cVEDA and IMAGEN factors match that of ABCD factors. With this, we assume that each factor in the three cohorts has its own distribution (PDF), but 90% of samples fall into similar range. The selection of 90% is an empirical choice, subject to change for different problems. The data harmonization was applied to anxiety/depression scores (cVEDA), ELS scores (cVEDA and IMAGEN), school risk scores(IMAGEN), air pollution scores(cVEDA), family conflict scores(cVEDA), and household income scores(cVEDA).

#### Analyzing effects of environmental factors using linear models

The impact of environmental factors on anxiety/depression in each cohort was analyzed using different methods including Linear Mixed models (LMMs) for each factor, and Recursive Feature Elimination (RFE) with linear regression as well as Least Absolute Shrinkage Selector Operator (LASSO) regression for the combination of factors.

In the case of LMMs, each of the individual environment factors was tested separately for all three cohorts. For the ABCD cohort, we tested the LMMs with sex as fixed effects, and family and site were considered as nested random effects. For LMMs implementation on cVEDA and IMAGEN cohorts, sex was considered a fixed effect, however, only site was considered a random effect because we had independent samples for both cohorts. For all tests, Bonferroni multiple comparison corrections were applied.

We also used RFE with linear regression to find the important environmental factors for the prediction of the anxiety/depression score for all three cohorts. In RFE, the importance of each feature in the model is calculated and ranked in order, and the feature with the least importance is removed iteratively based on evaluation metrics such as root mean squared error, accuracy, etc. In our case, the anxiety/depression score was used as the dependent variable for all three cohorts. The independent variables were nine environmental factors (including sex) for ABCD, six environmental factors (including sex) for cVEDA, and four environmental factors (including sex) for IMAGEN. For all three cohorts, the data were standardized and divided into training and testing sets (70/30), and 10-fold internal cross-validation was performed on the training data to find the best features. Using the best features from the internal cross-validation, the final model was trained using all training data and tested on the remaining 30% of testing data, and the explained variance (R2) was estimated and reported.

Along with RFE, we further validated the effect of the environmental factors using LASSO Regression. LASSO regression is a very popular regularization-based feature selection method in which the less important features are penalized by making the respective coefficients zero, and thereby eliminating them completely. The cost function for Lasso regression is represented as:

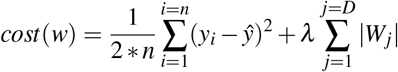

Here, *λ* is a parameter chosen by the internal cross-validation to decide how aggressive the regularization is performed (how sparse the feature space is). In this way, lasso regression removes the insignificant variables from the model. The independent variables used for LASSO models for the three cohorts were exactly the same to those in RFE models. So is the training and testing strategies with 70/30 splits and a 10-fold cross-validation on the training data to determine the regularization parameter (*λ*). The maximum explained variance was estimated on the test data.

#### Genome wide association study (GWAS) for each cohort

A univariate LMM was used to test the genome-wide association through the software package: genome-wide efficient mixed-model association algorithm (GEMMA)^7^. We estimated the relatedness matrix based on SNPs using GEMMA to account for the relatedness between samples for all three cohorts. The anxiety/depression score was used as the phenotype. For ABCD, covariates used were the significant environmental factors identified in the previous LMM test along with the 10 eigenvectors of genomic SNP data that represent the population stratification on ABCD data and the relatedness matrix of ABCD samples (random effect). Similarly, for cVEDA, covariates used were the significant environmental factors along with the 10 eigenvectors that represent the population stratification on cVEDA, age, and relatedness matrix of cVEDA samples (random effect). Finally, for IMAGEN covariates used were the significant environmental factors, 10 eigenvectors, and the relatedness matrix of IMAGEN samples (random effect). Merging the subjects with both the genetic data and the environmental factors available resulted in 7598 subjects and 8,367,466 SNPs for ABCD, 585 subjects and 4,472,935 SNPs for cVEDA, and 1580 subjects and 8,775,504 SNPs for IMAGEN respectively. As the phenotype(anxiety/depression score) for all three cohorts was not normally distributed, the rank-based inverse normal transformation was used to transform the dependent variable before testing for association using linear mixed models using GEMMA.

#### Meta analysis and Mega analysis

Both meta- and mega-analyses on genetic associations were performed to test the consistency of risk variants. We found 3,333,270 SNPs to be common across all three cohorts. For the meta-analysis, we applied the random effects model (RE2)^34^ from METASOFT on the results of individual GWAS performed for the three cohorts. RE2 model assumes different effect sizes across cohorts which are against a consist zero mean distribution under the null hypothesis.

Mega-analysis was performed by combining all three cohorts’ data together and performing a genome-wide association analysis using GEMMA. The covariates included the relatedness matrix and the 10 eigenvectors computed from the combined genomic data, the common environmental factors that had consistent, significant effects across all three cohorts, as well as age and cohorts. Age was coded as two groups (1 for 6-11 age range, 2 for 12-17 age range), since the ABCD cohort has an age range of 9-10, cVEDA cohort has an age range of 6-11, and 12-17, and IMAGEN has 14. The cohort was coded as two dummy variables.

#### Genomic risk loci and Gene mapping

Functional annotation was performed on SNP results from meta and mega-analyses results with FUMA^35^, an online platform for the functional mapping of genetic variants. We first defined ‘independent significant SNPs’ as those surpassing a predefined threshold P value (5E-06) and showing moderate to low linkage disequilibrium (r2 < 0.6). We further defined ‘lead SNPs’ as the subset of independent SNPs (r2 < 0.1). Genomic risk loci were identified by merging LD blocks of independent significant SNPs that have close physical positions (< 250 kb). All LD information was calculated from the 1000G phase3 ALL population. More details about LD clump can be found in FUMA website (https://fuma.ctglab.nl/tutorial). Genes involved in each genomic risk loci were mapped from SNPs using three strategies in FUMA. First, position mapping was based on the physical distances (within a 10 kb window) from SNPs to known protein-coding genes in the human reference assembly (GRCh38). The second strategy, expression quantitative trait loci (eQTL) mapping, used BrainEAC^36^ (11 brain tissues) and GTEx v8 Brain^37^ (13 tissues) eQTLs information to map SNPs to genes (i.e., where the expression of the gene is associated with allelic variation at the SNP, and the association survives false discovery rate (FDR) of 0.05). The third strategy, chromatin interaction mapping, mapped SNPs to the promoter regions of genes based on significant chromatin interactions. This type of mapping was a 3D DNA interaction between the SNP region and a gene region, without a distance boundary. FUMA currently contains Hi-C data for 21 tissue/cell types^38^. More details can be found in FUMA^35^.

#### Gene set and tissue specificity enrichment analyses

To explore if anxiety/depression associated mutations were enriched in specific human tissues, we performed tissue enrichment analysis for both meta-analysis and mega-analysis results by using MAGMA functions implemented in FUMA software. Briefly, gene expression data of different human tissues (RNA sequencing data from the GTEx consortium) were used to identify the genes that were differentially expressed in a specific tissue. Based on the individual SNPs association values, MAGMA quantifies the degree of association between a gene and anxiety/depression (i.e., obtain a gene-level P value) by using a multiple linear principal component regression models. MAGMA then tests if genes’ associated with anxiety/depression were enriched in the specifically expressed genes in a specific tissue. More detailed information about tissue enrichment analysis can be found on FUMA website (https://fuma.ctglab.nl/).

The common genes mapped from both meta-analysis and mega-analysis were selected to further investigate functional annotation using the GENE2FUNC procedure in FUMA. This procedure provides hypergeometric tests of enrichment in MSigDB gene sets^39^, including BioCarta, KEGG, Reactome, and Gene Oncology (GO). The P values for gene set enrichment analyses were adjusted by the Benjamini–Hochberg method. The threshold of the adjusted P-value was 0.05. The minimum number of input genes overlapping with a tested gene set to be reported as significant was two. Furthermore, the common mapped genes were also tested for enrichment in specific human tissues by performing tissue enrichment analysis in FUMA, where RNA sequencing data from the GTEx v8: 54 tissue types and GTEX v8: 30 general tissue types^40^ were used.

#### Determining the significance of Polygenic Risk Score (PRS)

The cumulative effect of genetics was obtained by PRS using the software PRS-CS^41^. PRS-CS uses the Bayesian regression framework that infers posterior SNP effect sizes under continuous shrinkage (CS) priors based on GWAS summary statistics and an external LD reference panel. Three LD reference panels were used: AMR(American) reference for ABCD, EUR(European) reference for IMAGEN and SAS(South Asian) reference for the CVEDA cohort. For GWAS summary statistics, we compared our own GWAS results with that from recently reported large sample GWAS on depression. Specifically, the summary statistics of GWAS for MDD from a large study conducted in 2019 with 246,363 cases and 561,190 controls from Europe and the United States^22^ were applied to the ABCD and IMAGEN cohorts, compared with our own GWAS summary statistics. For cVEDA cohort, we downloaded a large-scale GWAS for MDD performed on East Asian ancestry individuals^42^ with 15,771 cases and 178,777 controls. When using our own GWAS results and avoiding bias, we used IMAGEN GWAS results for ABCD cohort and ABCD GWAS results for IMAGEN and cVEDA cohorts. The significance of the generated PRS for each cohort was determined using a linear mixed-effect regression model to predict the anxiety and depression scores. For ABCD cohort, the model also included sex as a fixed effect covariate, and site and family as nested random effect covariates. For cVEDA cohort, the model also included sex and age as fixed effect covariates and site as a random effect covariate. For the IMAGEN cohort, only sex was included as a fixed effect and site as a random effect. Furthermore, we also tested the change in total variation explained by adding the PRS score as an additional fixed effect on the linear models that we used to analyze the effects of environmental factors for each cohort.

## Results

### Significant effect of environmental factors on the anxiety/depression score

The data harmonization was performed by comparing the CDF and performing gamma transformation on the anxiety/depression scores and some environmental factors of cVEDA and IMAGEN to match data from ABCD. As an illustration, Figure 1 shows the CDF of the anxiety/depression scores and ELS scores of the three cohorts before and after data harmonization. Other environmental factors’ CDF plots and parameters of gamma transformation can be in Supplementary files.

**Figure 1.**
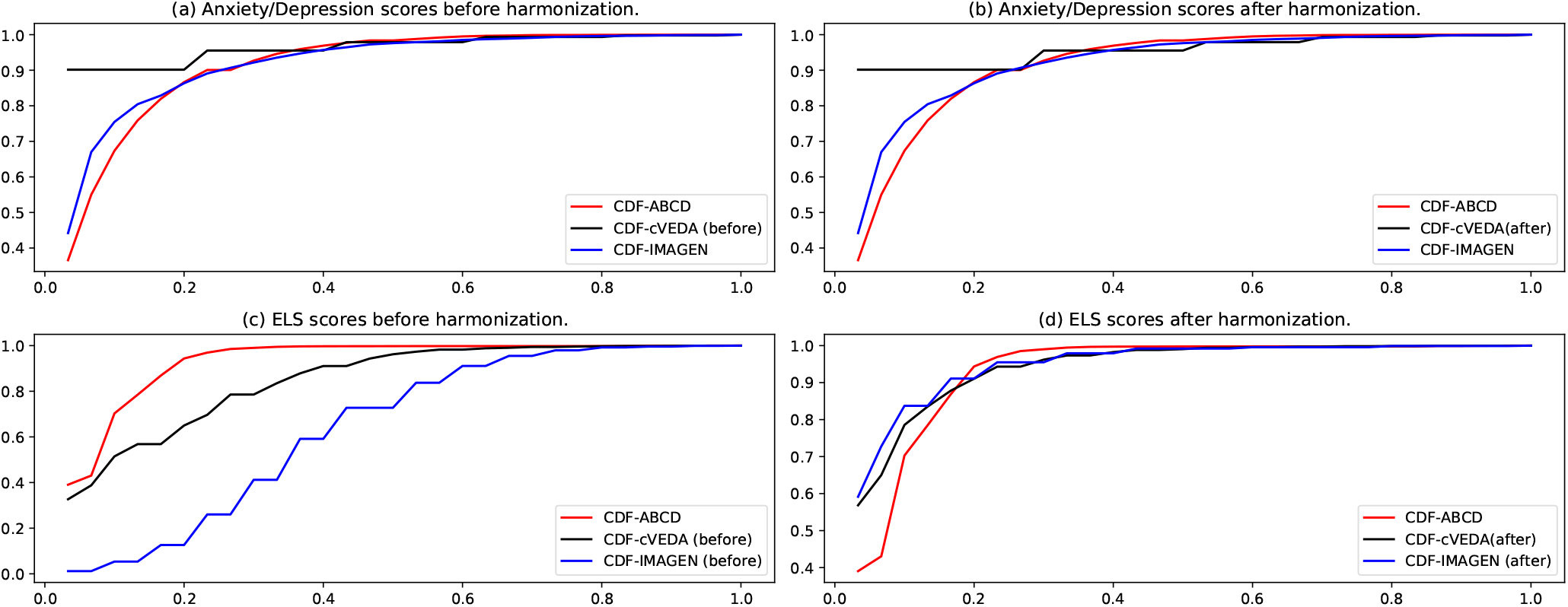
Data Harmonization. a) Anxiety/depression scores of ABCD, cVEDA and IMAGEN before applying gamma transformation. b) Anxiety/depression scores of ABCD, cVEDA and IMAGEN after applying gamma transformation (gamma=1.75x for cVEDA). No transformation needed for IMAGEN. c) ELS scores of ABCD, cVEDA and IMAGEN before applying gamma transformation. d) ELS scores of of ABCD, cVEDA and IMAGEN after applying gamma transformation (gamma=1.75x for cVEDA, 3.5x for IMAGEN)

With harmonized data and using three linear types of models (LMM, RFE and LASSO), we identified seven factors (environmental factors and sex) in ABCD cohort, four factors (including sex) in cVEDA, and three factors (including sex) in the IMAGEN cohort that were significantly related and contribute to the anxiety/depression score. The results of LMM for each cohort are presented in Table 1. ELS has the most significant effect across all three cohorts with effect sizes from a beta value of 0.304 to 0.424, where increasing ELS scores are associated with increasing anxiety/depression scores. Since we have harmonized data, the beta values in LMM models can be directly compared. The next significant and consistent factor is school risk with p values ranging from 6.02e-06 to 1.73e-37, and effects ranging from -0.081 to -0.182, indicating a better school environment leading to decrease anxiety/depression scores. Family conflict was found significantly affecting anxiety/depression in ABCD and cVEDA cohorts, but not in the IMAGEN cohort.

**Table 1.**
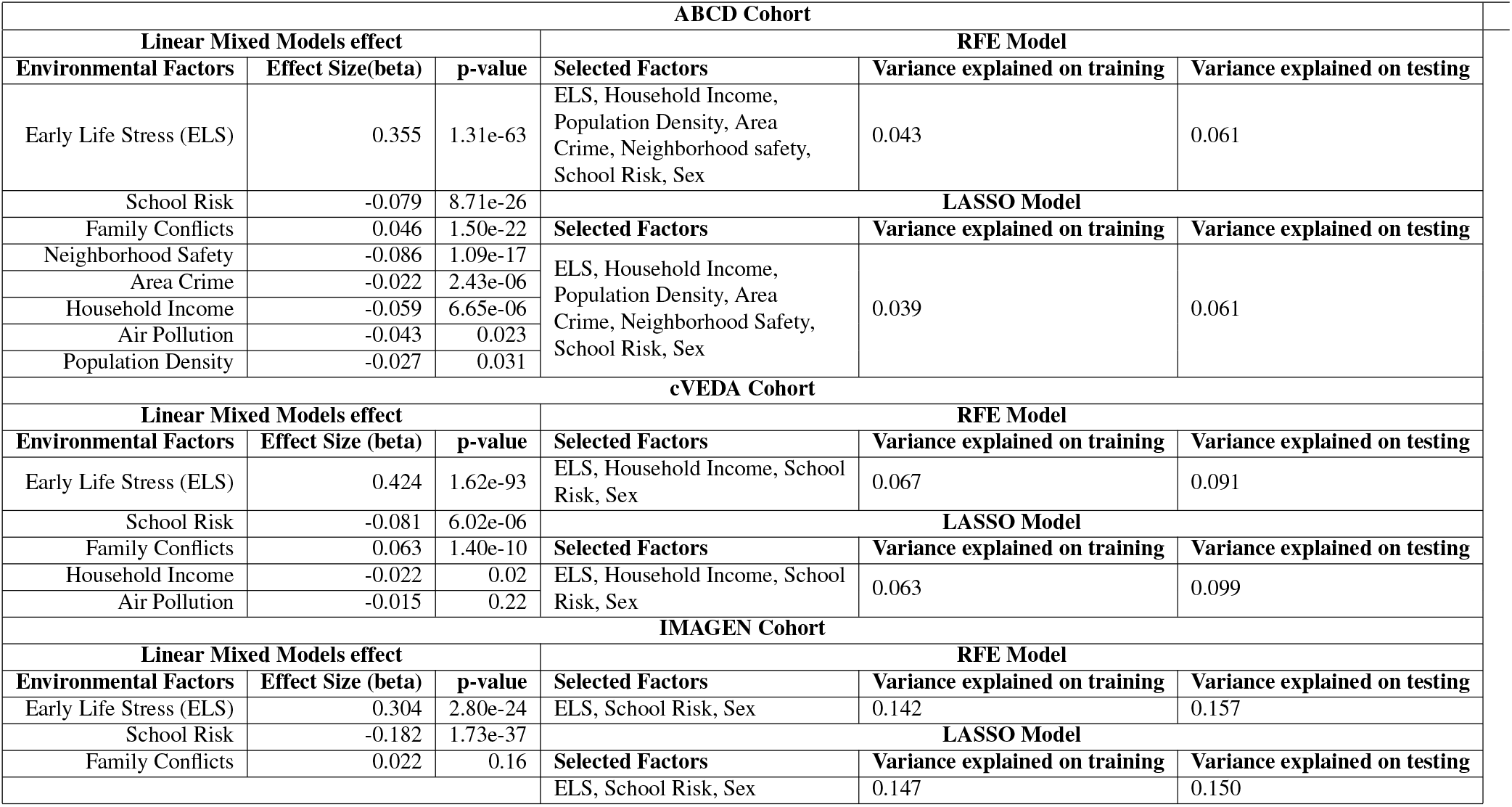
Output of RFE and LASSO models along with the individual environmental factors effect using Linear Mixed Models (LMMs).

**Table 2.**
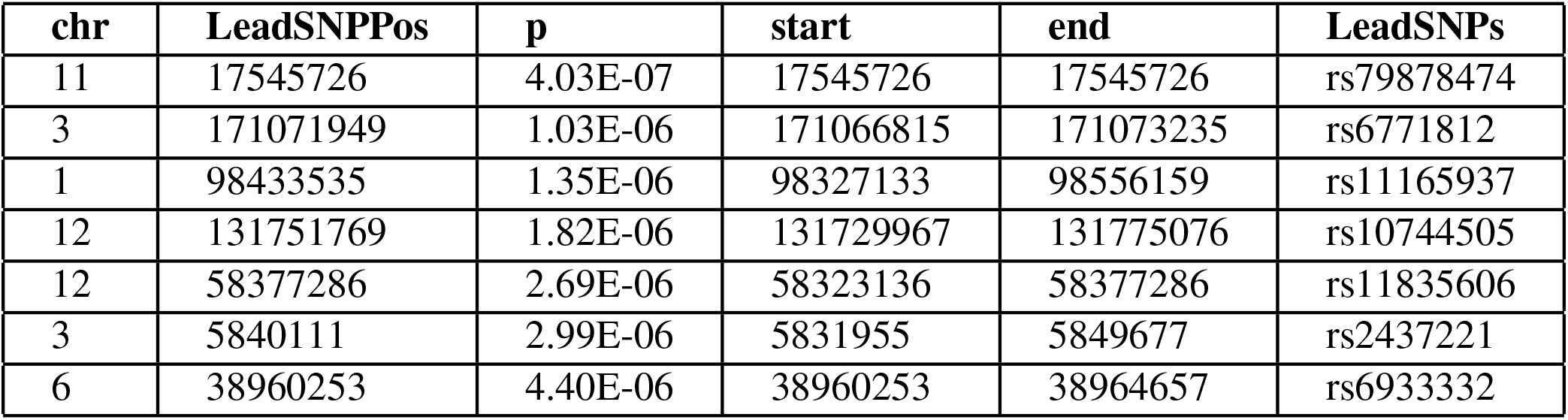
Identification of independent loci from mega-analysis GWAS using FUMA

| chr | LeadSNPPos | p | start | end | LeadSNPs |
| --- | --- | --- | --- | --- | --- |
| 11 | 17545726 | 4.03E-07 | 17545726 | 17545726 | rs79878474 |
| 3 | 171071949 | 1.03E-06 | 171066815 | 171073235 | rs6771812 |
| 1 | 98433535 | 1.35E-06 | 98327133 | 98556159 | rs11165937 |
| 12 | 131751769 | 1.82E-06 | 131729967 | 131775076 | rs10744505 |
| 12 | 58377286 | 2.69E-06 | 58323136 | 58377286 | rs11835606 |
| 3 | 5840111 | 2.99E-06 | 5831955 | 5849677 | rs2437221 |
| 6 | 38960253 | 4.40E-06 | 38960253 | 38964657 | rs6933332 |

**Table 3.** Identification of independent loci from meta-analysis GWAS using FUMA

| chr | LeadSNPPos | p | start | end | LeadSNPs |
| --- | --- | --- | --- | --- | --- |
| 11 | 17545726 | 1.13E-06 | 17545726 | 17545726 | rs79878474 |
| 3 | 171071949 | 1.24E-06 | 171066876 | 171073235 | rs6771812 |
| 2 | 38034558 | 1.25E-06 | 38031918 | 38034558 | rs6755353 |
| 6 | 38960253 | 2.86E-06 | 38960253 | 38964657 | rs6933332 |
| 3 | 80388728 | 3.49E-06 | 80388728 | 80493313 | rs6764488 |
| 7 | 95705989 | 3.75E-06 | 95705989 | 95711226 | rs756859 |
| 10 | 115522548 | 4.44E-06 | 115522548 | 115522548 | rs2900993 |

RFE and LASSO models selected the optimal number of features which was seven for ABCD, four for IMAGEN and three on IMAGEN. In RFE models the maximum explained variance on the remaining 30% of the test data was 6.1% for ABCD, 9.1% for cVEDA, and 15.7% for IMAGEN. In the LASSO regression model, the regularization parameter(lambda) estimated using the 10-fold cross-validation was 0.006 for ABCD, 0.021 for cVEDA, and 0.006 for IMAGEN (Figures in Supplementary files). The maximum variance explained by LASSO on 30% of the test data was 6.1% for ABCD, 9.9% for cVEDA, and 15% for IMAGEN.

For ABCD cohort, both RFE and LASSO models selected sex and six environmental factors (ELS, household income, population density, area crime, neighborhood safety, and school risk), and ignored the two factors (air pollution and family conflicts) considering their contribution not significant. For the cVEDA cohort, sex and three environmental factors (ELS, household income and school risk) were selected, and two factors (air pollution and family conflicts) were considered not contributing. Finally, for IMAGEN cohort, sex and two environmental factors (ELS, School Risk) were considered to have a significant contributions, whereas family conflicts factor was not considered contributing. Thus, these selected factors were used as covariates in the following GWAS analyses for each cohort, and mega-analysis of GWAS used common significant contributors including ELS, school risk, and sex, in addition to age and cohort.

### Result of mega-analysis and meta-analysis on SNPs and genes

The genomic inflation factor (*λ*) in the QQ Plot for mega- and meta-analyses was 1.012 and 1.003 respectively, indicating no systemic bias in the analyses. Although, mega-analysis and meta-analysis did not find any SNPs to be significantly associated (p<5e-08) with anxiety/depression score, we found many promising SNPs with p values less than p<5e-06. The MEGA analysis found 16 SNPs (Supplementary Table 5) to be promising with the most promising SNP as rs79878474, with p= 4.03e-07. The META analysis found 11 SNPs (Supplementary Table 4) to be promising with the same most promising SNP being rs79878474 (p=1.13E-06). In fact, the top three promising SNPs from mega-analysis (rs79878474, rs67861307, and rs6771812) were the same ones from meta-analysis. The complete set of results of mega-analysis and meta-analysis as well as each individual cohort’s analyses, and the corresponding Manhattan and QQ Plots can be found in Supplementary Files.

We further used FUMA to identify independent risk loci in the promising SNPs from meta-analysis and mega-analysis respectively. 7 independent risk loci were identified from mega-analysis (Table 2), mapped to 7 lead SNPs, 182 candidate SNPs, and 44 genes. Similarly, 7 independent risk loci were identified from the meta-analysis (Table 3), mapped to 7 lead SNPs, 82 candidate SNPs, and 58 genes. There are three common independent risk loci between meta- and mega-analyses: chr11:17545726, chr3:171071949, and chr6:38960253.

### Results of gene set and tissue enrichment analyses

For the gene set enrichment analyses, we selected 20 common genes (Supplementary Table 13) from meta-analysis mapped genes and mega-analysis mapped genes. Among a total of 10,678 gene sets, 49 gene sets were considered to be statistically significant (Supplementary Table 10). They are grouped into three categories (positional, functional and GWAS Catalog) and consolidated with shared overlapped genes as listed in Table 4. The positional gene sets chr11p15 (p=8.35E-14) and chr3q36 (p=3.33E-07) had the lowest p-values. The GO biological processes gene sets with the lowest p-values include regulation of insulin/hormone/peptide secretion, and regulation of potassium channel. The GO cellular component gene sets with the lowest p-values include potassium channel complex, synapse, and axolemma. Three significant gene sets from the GWAS catalog were systolic blood pressure x alcohol consumption interaction, body mass index, and night sleep phenotypes. Reactome and KEGG databases identified similar related gene sets(Supplementary Table 10).

**Table 4.**
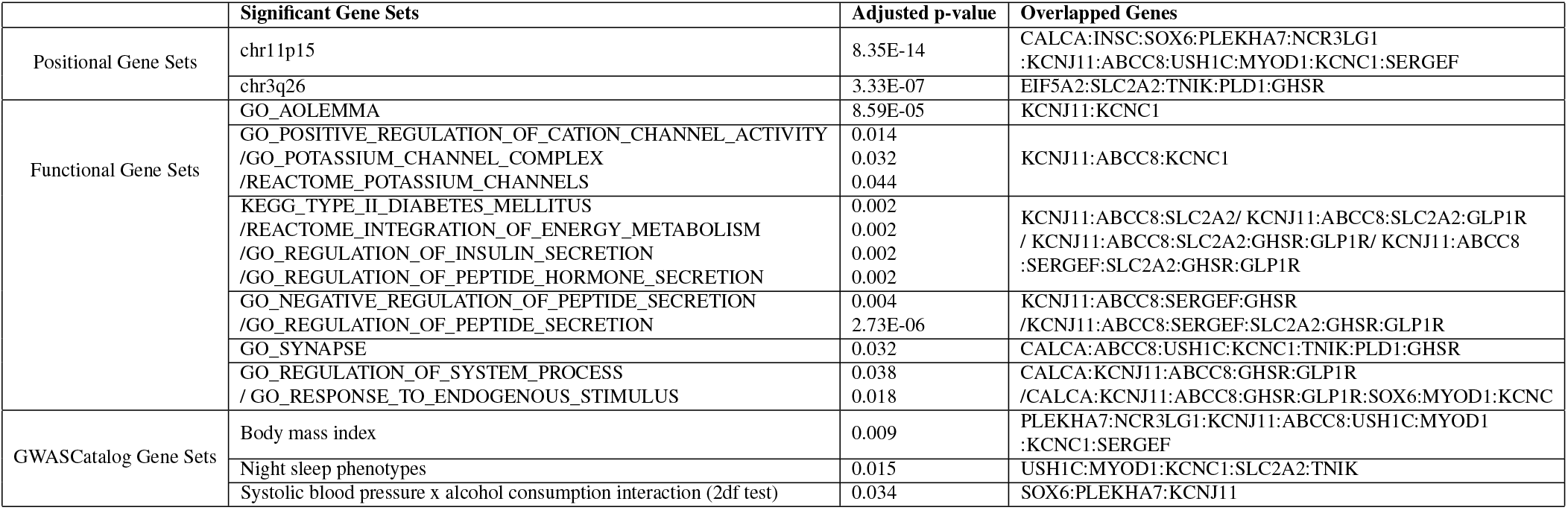
Identification of gene and gene sets associated with anxiety/depression using FUMA

For the tissue enrichment analysis, when tested individually for meta- and mega-analysis results using MAGMA, both meta-and mega analyses results showed an elevated enrichment in the brain cerebellum with uncorrected p value of 0.007 and 0.003, respectively, tested for 53 tissue types (Supplementary Table 14 and 15), although not passing multiple comparison correction. In contrast, when performing the tissue enrichment test for 20 common genes using GENE2FUNC in FUMA, tissues in the small intestine showed significant enrichment with an adjusted p-value of 0.04 tested for 53 tissue types. See Supplementary files for detailed results on tissue expression analysis using FUMA.

### Significance of PRS

Analyses of the PRS on the anxiety/depression for the three cohorts showed that the PRS score was only statistically significantly associated with the anxiety/depression in ABCD cohort, and not significant in cVEDA and IMAGEN cohorts. In ABCD cohort, both PRS scores computed using either our own GWAS summary statistics of IMAGEN cohort or recently reported large scale GWAS statistics showed significant p-values (p < 6.23e-03 and p < 4.56e-14 respectively). However, the percentage of variation explained were small, i.e., the total variance explained remained approximately unchanged after the addition of PRS as an independent variable along with significant environmental factors.

## Discussion

In this study we tested the effects of different environmental factors on anxiety and depression in children and adolescents across three diverse cohorts, and further studied genetic variants associated with the consideration of environmental factors. The three cohorts in three different continents are under different environmental backgrounds at a large scale. Yet, we hypothesize that each environmental factor could have a consistent effect, maybe not with the same level of strength, on anxiety and depression during the development across diverse backgrounds. While the three cohorts used different environmental and anxiety/depression measures, we conducted data harmonization to allow comparison, meta-analysis, and mega-analysis of the results. In other words, each data set after harmonization is from the same scale but with its own distributive characteristics to make the resultant findings comparable. The maximum variance explained by the environmental factors was in the range of 6.1 % to 15%. Note that ELS and school risk were consistently selected by RFE and LASSO, with the explained variance being largely comparable across the three cohorts, lending support for the effectiveness of data harmonization. It is interesting to note that school risk had a significant consistent effect in addition to ELS. This implies that the way children are treated and behaved in school will have a significant impact on their mental health, and a better environment in school might help to reduce anxiety/depression. Meanwhile, family conflicts is highly correlated to ELS, such that this factor was eliminated by RFE and LASSO due to not providing additional information^26^. It is noted that family conflicts were not significant in the IMAGEN cohort even when tested individually. Looking at the original data distribution before harmonization, family conflicts from IMAGEN presented very different CDF as compared to other cohorts (ABCD and cVEDA). In the case of IMAGEN, half of the population reported an incidence of family conflicts below 0.65, while half of the ABCD subjects reported an incidence below 0.20 in a scale of 0 to 1. This could be due to more willingness to report the incidence of family conflicts in the case of IMAGEN, which might contribute to inconsistent effects.

Although mega- and meta-analyses both incorporate effects from three cohorts, mega-analysis assumes one homogeneous effect size from all three cohorts, while random-effect meta-analysis we implemented^34^ allows different effect size across cohorts. Thus, we expect some level of consistence and differences between meta- and mega-analyses results. Both analyses identified the same three top risk SNPs with the most promising SNP as rs79878474 with p value of 4.03E-7 (mega-analyses). This SNP is located in USH1C gene which is highly expressed in the brain, particularly in the spinal cord, following small intestine based on GTEx V8 (https://gtexportal.org/home/). Functionally, gene USH1C encodes a scaffold protein that functions in the assembly of Usher protein complexes and mutation of USH1C is known to be involved Usher syndrome type 1C and sensorineural deafness^43^. The other two top SNPs are in the TNIK gene (TRAF2 and NCK interacting kinase), which is also highly expressed in brain and has been shown to regulate neurite development^44^, and mutations involved with an autosomal recessive form of cognitive disability^45^. But how these SNPs and genes related to anxiety/depression during development warrants further investigation. In general, mega-analysis is preferred compared to meta-analysis under the same homogeneous condition as showed by a recent empirical comparison where under the same condition the mega-analysis produces lower standard errors and narrower confidence intervals than the meta-analysis^46^. Nevertheless, the mega-analysis requires high agreement on the variables collected from different sites; the same variables and the same assessments are used from all sites. As in our study, after data harmonization to make the mega-analysis possible as the variables were measured differently in each site, mega-analysis only considered three common contributing factors (ELS, school risk, and sex), while random-effect meta-analysis was able to control for specific environmental factors’ effect for each cohort separately, and allows cohort-specific genetic effect size. It is not surprising to see some differences in the results of meta- and meta-analyses. Given both meta- and mega-analyses have strengths and limitations, our study focuses on common independent risk loci and commonly indicated genes from both analyses.

FUMA identified three common independent risk loci with lead SNPs as rs79878474, rs6771812, rs6933332, and 20 common mapped genes between meta- and mega-analyses. The subsequent gene set analysis found 49 statistically significant gene sets with the most significant being chr11p15 and chr3q26 positional gene sets. Enriched gene sets from GO, KEGG and Reactome database are categorized based on similar overlapping genes, including functions related to potassium channels, insulin/energy metabolism/peptide secretion, and synapse and system process. We want to highlight potassium channel regulation here with genes KCNJ11, KCNC1 and ABCC8. Potassium (K+) channels locate in cell membranes and control the transportation of K+ ions efflux from and the influx into cells. This superfamily can be divided into many structural classes and located in different tissue types^47^, but most classes are prominent in ventricular tissue to regulate cardiac function, and in the brain (neurons, soma, dendrites, and axons) to influence neural activities^48^. KCNC1 is highly and exclusively expressed in the cerebellum based on GTEx and encodes member 1, subfamily C of integral membrane proteins that mediate the voltage-dependent potassium ion permeability of excitable membranes. This protein is the key to K+ voltage-dependent channel Kv3^48,49^. Kv3 channels regulate neurotransmitter release^50^, particularly affecting the high-frequency firing of neuron^51^ including circadian rhythms in the suprachiasmatic nucleus of the hypothalamus^52^. Alterations of Kv3 channels’ properties can cause severe neurological disorders like epilepsy and broad phenotypic spectrum including developmental delay^53^, schizophrenia^54^, and depression^55^. Recent animal and cell line studies have strengthened the connection between the Kv3 channel and depression. Mice with a reduced level of Kv3.1 presented vulnerability to depressive behavior, whereas up-regulation of Kv3.1 or acute activation of Kv3.1 induced resilience to depression^56^. A commonly used antidepressant drug, Fluoxetine, acts on Kv3 channels to affect Kv3.1b expression and serotonin secretion in a serotonergic cell line^57^, and another similar drug Vortioxetine inhibits delayed-rectifier K+ current caused by Kv3 channels activity in pituitary GH3 cells^58^. KCNJ11 is highly expressed in the cerebellum (the second highest besides muscle) and encodes an integral membrane protein that is the key to an inward-rectifier potassium channel, the Kir6.2 subunit of ATP-sensitive potassium channel. Kir6.2 channel is known to play an important role in modulating insulin secretion^48^, and also plays a role in stress adaptation^59,60^, as well as possibly part of the mechanism for anti-depression effect^60,61^. ABCC8 is expressed mainly in cerebellum followed by the frontal cortex pituitary, and pancreas. Functionally it modulates the SUR subunit of ATP-sensitive potassium channel which plays a key role in mediating glucose-stimulated insulin secretion. Recently new studies have linked insulin resistance with risk for depression and anxiety^62–64^. Our findings further strength this association by discovery of both potassium channel gene sets and insulin secretion gene sets in association of depression and anxiety score. Our findings suggest that the Kv3, Kir 6.2,and SUR subunit of potassium channels may be important targets for anti-depression treatment.

The tissue enrichment analysis using MAGMA for either meta- or mega-analysis results showed an elevated expression enrichment in the brain, more specifically, in the cerebellum region of the brain. Common genes between meta- and mega-analyses showed significant tissue enrichment in the small intestine by GENE2FUNC in FUMA package. The importance of cerebellum is supported by expression of key genes in the potassium channels as discussed above. Gene USH1C has the highest expression in small intestine as well as the spinal cord and other areas of the brain, while genes INSC, SOX6, PLEKHA7, SLC2A2, and TNIK are expressed in small intestine. The relation between small intestine and depression/anxiety has long been hinted particularly by brain-gut connections^65,66^. Our results emphasize small intestine and cerebellum in relation to depression and anxiety, which is not totally surprising but needs further in-depth investigation.

The significant association between the PRS and anxiety/depression score was only observed in the ABCD cohort, not in IMAGEN and cVEDA cohorts. We believe that the most likely reason is small sizes and small effect size, as in the ABCD cohort even though the variance explained by PRS is not big but with large sample sizes we could detect a significant PRS contribution. Interestingly, both PRS scores computed using our own GWAS or the downloaded large sample GWAS summary statistics showed significant contribution to anxiety/depression in the ABCD cohort, lending support to the validity of our GWAS analyses using relatively small samples but with careful controlling for the environmental factors.

To summarize, our findings show that there is a consistent environmental influence, particularly ELS and school risks, on anxiety and depression in children and adolescents across continents. Further research into the genetic susceptibility from meta- and mega-analyses highlights mutations and gene sets in chromosome 11 p15 region (chr11p15), and gene sets in potassium channels (Kv3, Kir 6.2, and SUR subunit) which are highly, if not exclusively, expressed in the brain cerebellum, were enriched for association with anxiety and depression. These findings, in line with literature about the potassium channel’s involvement in (anti)depression, and insulin secretion association with depression, motivate further investigation on how Kv3, Kir 6.2, SUR potassium channels in the cerebellum regulate anxiety and depression. For future work, we can incorporate the brain imaging data of subjects used in this study, focusing on the cerebellum region, and test brain structural and functional associations with anxiety and depression, and the effects of environmental and genetic influence on the brain to further validate the current results. Furthermore, we will also study dynamic changes of genetic and environmental effects on the brain and depression using data from the same participants in coming years, to keep track of the rate of mental changes and the effects as they develop.

## Supporting information

Supplementary Files - Description

## Data Availability

Data used in the preparation of this article were obtained from the Adolescent Brain Cognitive Development (ABCD) Study (https://abcdstudy.org), held in the NIMH Data Archive (NDA). Another set of data used is cVEDA, which is jointly funded by the Indian Council for Medical Research (ICMR/MRC/3/M/2015-NCD-I) and the Newton Grant from the Medical Research Council(MR/N000390/1), United Kingdom. Furthermore, the IMAGEN study was also used which receives research funding from the European Community Sixth Framework Programme (LSHM-CT-2007-037286).

https://cveda-project.org/

https://pubmed.ncbi.nlm.nih.gov/32601453/

## Acknowledgements

This study was funded part by NIH grant R01DA049238 and NSF grant 2112455. We thank the Adolescent Brain Cognitive Development (ABCD), Consortium on Vulnerability to Externalizing Disorders and Addictions (cVEDA) and IMAGEN participants and their families for their time and dedication to this project. Data used in the preparation of this article were obtained from the Adolescent Brain Cognitive Development (ABCD) Study (https://abcdstudy.org), held in the NIMH Data Archive (NDA). This is a multisite, longitudinal study designed to recruit more than 10,000 children aged 9-10 and follow them over 10 years into early adulthood. The ABCD Study is supported by the National Institutes of Health and additional federal partners under award numbers U01DA041048, U01DA050989, U01DA051016, U01DA041022, U01DA051018, U01DA051037, U01DA050987, U01DA041174, U01DA041106, U01DA041117, U01DA041028, U01DA041134, U01DA050988, U01DA051039, U01DA041156, U01DA041025, U01DA041120, U01DA051038, U01DA041148, U01DA041093, U01DA041089, U24DA041123, U24DA041147. A full list of supporters is available at https://abcdstudy.org/federal-partners.html. A listing of participating sites and a complete listing of the study investigators can be found at https://abcdstudy.org/consortium_members. ABCD consortium investigators designed and implemented the study and/or provided data but did not necessarily participate in analysis or writing of this report. This manuscript reflects the views of the authors and may not reflect the opinions or views of the NIH or ABCD consortium investigators. cVEDA is jointly funded by the Indian Council for Medical Research (ICMR/MRC/3/M/2015-NCD-I) and the Newton Grant from the Medical Research Council(MR/N000390/1), United Kingdom. IMAGEN study receives research funding from the European Community’s Sixth Framework Programme (LSHM-CT-2007-037286). This paper reflects only the author’s views, and the community is not liable for any use that may be made of the information contained therein. We thank all families for their help with this study.

## Author contributions statement

B.T. conducted the primary data analysis, experiments, writing, and reviewing of the manuscript. J.C. was involved in data analysis, discussion of the analyses, and reviewing the manuscript. B.R., B.F., P.S, and R.S were involved in the discussion of the analyses and reviewing the manuscript. I.C and C.C provided the IMAGEN and cVEDA datasets to work with. B.H, J.M, and N.V. prepared the cVEDA dataset and reviewed the manuscript. N.P.B. assisted with the discussion and manuscript review. V.B. was involved in the preparation and providing access to cVEDA dataset. G.S. was involved in the design of the project, providing access to cVEDA and IMAGEN cohorts, and reviewing the manuscript. V.D.C was involved in the design of the project and the manuscript review. J.L. conceived of the original idea and was involved in the design, data analysis, and reviewing of the manuscript. All authors reviewed the manuscript.

## References

1. Merikangas, K. R. et al. Lifetime prevalence of mental disorders in u.s. adolescents: Results from the national comorbidity survey replication–adolescent supplement (NCS-a). J. Am. Acad. Child & Adolesc. Psychiatry 49, 980–989, DOI: 10.1016/j.jaac.2010.05.017 (2010).

2. Ghandour, R. M. et al. Prevalence and treatment of depression, anxiety, and conduct problems in US children. The J. Pediatr. 206, 256–267.e3, DOI: 10.1016/j.jpeds.2018.09.021 (2019).

3. Kessler, R. C., Merikangas, K. R. & Wang, P. S. Prevalence, comorbidity, and service utilization for mood disorders in the united states at the beginning of the twenty-first century. Annu. Rev. Clin. Psychol. 3, 137–158, DOI: 10.1146/annurev.clinpsy.3.022806.091444 (2007).

4. Kaufman, J. & Charney, D. Comorbidity of mood and anxiety disorders. Depress. Anxiety 12, 69–76, DOI: 10.1002/1520-6394(2000)12:1+<69::aid-da9>3.0.co;2-k (2000).

5. Hasin, D. S., Goodwin, R. D., Stinson, F. S. & Grant, B. F. Epidemiology of major depressive disorder. Arch. Gen. Psychiatry 62, 1097, DOI: 10.1001/archpsyc.62.10.1097 (2005).

6. Johannessen, E. L., Andersson, H. W., Bjørngaard, J. H. & Pape, K. Anxiety and depression symptoms and alcohol use among adolescents - a cross sectional study of norwegian secondary school students. BMC Public Heal. 17, DOI: 10.1186/s12889-017-4389-2 (2017).

7. Zhou, X. & Stephens, M. Genome-wide efficient mixed-model analysis for association studies. Nat. Genet. 44, 821–824, DOI: 10.1038/ng.2310 (2012).

8. Brooks-Gunn, J. & Duncan, G. J. The effects of poverty on children. The Futur. Child. 7, 55, DOI: 10.2307/1602387 (1997).

9. McLeod, J. D. & Shanahan, M. J. Trajectories of poverty and children’s mental health. J. Heal. Soc. Behav. 37, 207, DOI: 10.2307/2137292 (1996).

10. Repetti, R. L., Taylor, S. E. & Seeman, T. E. Risky families: Family social environments and the mental and physical health of offspring. Psychol. Bull. 128, 330–366, DOI: 10.1037/0033-2909.128.2.330 (2002).

11. Gilman, S. E., Kawachi, I., Fitzmaurice, G. M. & Buka, S. L. Family disruption in childhood and risk of adult depression. Am. J. Psychiatry 160, 939–946, DOI: 10.1176/appi.ajp.160.5.939 (2003).

12. Slopen, N., Koenen, K. C. & Kubzansky, L. D. Cumulative adversity in childhood and emergent risk factors for long-term health. The J. Pediatr. 164, 631–638.e2, DOI: 10.1016/j.jpeds.2013.11.003 (2014).

13. Widom, C. S., DuMont, K. & Czaja, S. J. A prospective investigation of major depressive disorder and comorbidity in abused and neglected children grown up. Arch. Gen. Psychiatry 64, 49, DOI: 10.1001/archpsyc.64.1.49 (2007).

14. Kessler, R. C. THE EFFECTS OF STRESSFUL LIFE EVENTS ON DEPRESSION. Annu. Rev. Psychol. 48, 191–214, DOI: 10.1146/annurev.psych.48.1.191 (1997).

15. Hammen, C. Stress and depression. Annu. Rev. Clin. Psychol. 1, 293–319, DOI: 10.1146/annurev.clinpsy.1.102803.143938 (2005).

16. Levey, D. F. et al. Reproducible genetic risk loci for anxiety: Results from ∼200,000 participants in the million veteran program. Am. J. Psychiatry 177, 223–232, DOI: 10.1176/appi.ajp.2019.19030256 (2020).

17. Otowa, T. et al. Meta-analysis of genome-wide association studies for panic disorder in the japanese population. Transl. Psychiatry 2, e186–e186, DOI: 10.1038/tp.2012.89 (2012).

18. Otowa, T. et al. Erratum: Meta-analysis of genome-wide association studies of anxiety disorders. Mol. Psychiatry 21, 1485–1485, DOI: 10.1038/mp.2016.11 (2016).

19. Levey, D. F. et al. Reproducible genetic risk loci for anxiety: Results from ∼200, 000 participants in the million veteran program. Am. J. Psychiatry 177, 223–232, DOI: 10.1176/appi.ajp.2019.19030256 (2020).

20. Purves, K. L. et al. A major role for common genetic variation in anxiety disorders. Mol. Psychiatry 25, 3292–3303, DOI: 10.1038/s41380-019-0559-1 (2019).

21. Sullivan, P. F., Neale, M. C. & Kendler, K. S. Genetic epidemiology of major depression: Review and meta-analysis. Am. J. Psychiatry 157, 1552–1562, DOI: 10.1176/appi.ajp.157.10.1552 (2000).

22. Howard, D. M. et al. Genome-wide meta-analysis of depression identifies 102 independent variants and highlights the importance of the prefrontal brain regions. Nat. Neurosci. 22, 343–352, DOI: 10.1038/s41593-018-0326-7 (2019).

23. Wray, N. R. et al. Genome-wide association analyses identify 44 risk variants and refine the genetic architecture of major depression. Nat. Genet. 50, 668–681, DOI: 10.1038/s41588-018-0090-3 (2018).

24. Forstner, A. J. et al. Genome-wide association study of panic disorder reveals genetic overlap with neuroticism and depression. Mol. Psychiatry 26, 4179–4190, DOI: 10.1038/s41380-019-0590-2 (2019).

25. Rice, F. Genetics of childhood and adolescent depression: insights into etiological heterogeneity and challenges for future genomic research. Genome Medicine 2, 68, DOI: 10.1186/gm189 (2010).

26. Thapaliya, B., Calhoun, V. D. & Liu, J. Environmental and genome-wide association study on children anxiety and depression. In 2021 IEEE International Conference on Bioinformatics and Biomedicine (BIBM) (IEEE, 2021).

27. Ghandour, R. M. et al. Prevalence and treatment of depression, anxiety, and conduct problems in US children. The J. Pediatr. 206, 256–267.e3, DOI: 10.1016/j.jpeds.2018.09.021 (2019).

28. Garavan, H. et al. Recruiting the ABCD sample: Design considerations and procedures. Dev. Cogn. Neurosci. 32, 16–22, DOI: 10.1016/j.dcn.2018.04.004 (2018).

29. Papdopoulos Orfanos, D. et al. c-veda dataset, DOI: 10.25720/VEDA-CMRH (2018).

30. Maričić, L. M. et al. The IMAGEN study: a decade of imaging genetics in adolescents. Mol. Psychiatry 25, 2648–2671, DOI: 10.1038/s41380-020-0822-5 (2020).

31. Taliun, D. et al. Sequencing of 53, 831 diverse genomes from the NHLBI TOPMed program. Nature 590, 290–299, DOI: 10.1038/s41586-021-03205-y (2021).

32. Das, S. et al. Next-generation genotype imputation service and methods. Nat. Genet. 48, 1284–1287, DOI: 10.1038/ng.3656 (2016).

33. Kent, W. J. et al. The human genome browser at UCSC. Genome Res. 12, 996–1006, DOI: 10.1101/gr.229102 (2002).

34. Lee, C. H., Eskin, E. & Han, B. Increasing the power of meta-analysis of genome-wide association studies to detect heterogeneous effects. Bioinformatics 33, i379–i388, DOI: 10.1093/bioinformatics/btx242 (2017).

35. Watanabe, K., Taskesen, E., van Bochoven, A. & Posthuma, D. Functional mapping and annotation of genetic associations with FUMA. Nat. Commun. 8, DOI: 10.1038/s41467-017-01261-5 (2017).

36. Ramasamy, A. et al. Genetic variability in the regulation of gene expression in ten regions of the human brain. Nat. Neurosci. 17, 1418–1428 (2014).

37. Genetic effects on gene expression across human tissues. Nature 550, 204–213, DOI: 10.1038/nature24277 (2017).

38. Schmitt, A. D. et al. A compendium of chromatin contact maps reveals spatially active regions in the human genome. Cell Reports 17, 2042–2059, DOI: 10.1016/j.celrep.2016.10.061 (2016).

39. Liberzon, A. et al. Molecular signatures database (MSigDB) 3.0. Bioinformatics 27, 1739–1740, DOI: 10.1093/bioinformatics/btr260 (2011).

40. Lonsdale, J. et al. The genotype-tissue expression (GTEx) project. Nat. Genet. 45, 580–585, DOI: 10.1038/ng.2653 (2013).

41. Ge, T., Chen, C.-Y., Ni, Y., Feng, Y.-C. A. & Smoller, J. W. Polygenic prediction via bayesian regression and continuous shrinkage priors. Nat. Commun. 10, DOI: 10.1038/s41467-019-09718-5 (2019).

42. Giannakopoulou, O. et al. The genetic architecture of depression in individuals of east asian ancestry. JAMA Psychiatry 78, 1258, DOI: 10.1001/jamapsychiatry.2021.2099 (2021).

43. Lentz, J. J. et al. Deafness and retinal degeneration in a novel USH1c knock-in mouse model. Dev. Neurobiol. 70, 253–267, DOI: 10.1002/dneu.20771 (2010).

44. Kawabe, H. et al. Regulation of rap2a by the ubiquitin ligase nedd4-1 controls neurite development. Neuron 65, 358–372, DOI: 10.1016/j.neuron.2010.01.007 (2010).

45. Anazi, S. et al. A null mutation in TNIK defines a novel locus for intellectual disability. Hum. Genet. 135, 773–778, DOI: 10.1007/s00439-016-1671-9 (2016).

46. Boedhoe, P. S. W. et al. An empirical comparison of meta-and mega-analysis with data from the ENIGMA obsessive-compulsive disorder working group. Front. Neuroinformatics 12, DOI: 10.3389/fninf.2018.00102 (2019).

47. Kuang, Q., Purhonen, P. & Hebert, H. Structure of potassium channels. Cell. Mol. Life Sci. 72, 3677–3693, DOI: 10.1007/s00018-015-1948-5 (2015).

48. González, C. et al. K+ channels: Function-structural overview, DOI: 10.1002/cphy.c110047 (2012).

49. Chi, G. et al. Cryo-EM structure of the human kv3.1 channel reveals gating control by the cytoplasmic t1 domain. Nat. Commun. 13, DOI: 10.1038/s41467-022-29594-w (2022).

50. Brooke, R. E., Moores, T. S., Morris, N. P., Parson, S. H. & Deuchars, J. Kv3 voltage-gated potassium channels regulate neurotransmitter release from mouse motor nerve terminals. Eur. J. Neurosci. 20, 3313–3321, DOI: 10.1111/j.1460-9568.2004.03730.x (2004).

51. Kaczmarek, L. K. & Zhang, Y. Kv3 channels: Enablers of rapid firing, neurotransmitter release, and neuronal endurance. Physiol. Rev. 97, 1431–1468, DOI: 10.1152/physrev.00002.2017 (2017).

52. Kudo, T., Loh, D. H., Kuljis, D., Constance, C. & Colwell, C. S. Fast delayed rectifier potassium current: Critical for input and output of the circadian system. J. Neurosci. 31, 2746–2755, DOI: 10.1523/jneurosci.5792-10.2011 (2011).

53. Park, J. et al. Kcnc1-related disorders: new de novo variants expand the phenotypic spectrum. Annals Clin. Transl. Neurol. 6, 1319–1326, DOI: 10.1002/acn3.50799 (2019).

54. Parekh, P. K. et al. Antimanic efficacy of a novel kv3 potassium channel modulator. Neuropsychopharmacology 43, 435–444, DOI: 10.1038/npp.2017.155 (2017).

55. Espinosa, F., Marks, G., Heintz, N. & Joho, R. H. Increased motor drive and sleep loss in mice lacking kv3-type potassium channels. Genes, Brain Behav. 3, 90–100, DOI: 10.1046/j.1601-183x.2003.00054.x (2004).

56. Medrihan, L. et al. Reduced kv3.1 activity in dentate gyrus parvalbumin cells induces vulnerability to depression. Biol. Psychiatry 88, 405–414, DOI: 10.1016/j.biopsych.2020.02.1179 (2020).

57. Tabka, H. et al. First evidence of kv3.1b potassium channel subtype expression during neuronal serotonergic 1c11 cell line development. Int. J. Mol. Sci. 21, 7175, DOI: 10.3390/ijms21197175 (2020).

58. Hsiao, H.-T., Wang, J. C.-F. & Wu, S.-N. Inhibitory effectiveness in delayed-rectifier potassium current caused by vortioxetine, known to be a novel antidepressant. Biomedicines 10, 1318, DOI: 10.3390/biomedicines10061318 (2022).

59. Zingman, L. V. et al. Kir6.2 is required for adaptation to stress. Proc. Natl. Acad. Sci. 99, 13278–13283, DOI: 10.1073/pnas.212315199 (2002).

60. Fan, Y., Kong, H., Ye, X., Ding, J. & Hu, G. ATP-sensitive potassium channels: uncovering novel targets for treating depression. Brain Struct. Funct. 221, 3111–3122, DOI: 10.1007/s00429-015-1090-z (2015).

61. Krzystanek, M., Surma, S., Pałasz, A., Romańczyk, M. & Krysta, K. Possible antidepressant effects of memantine—systematic review with a case study. Pharmaceuticals 14, 481, DOI: 10.3390/ph14050481 (2021).

62. Kleinridders, A. et al. Insulin resistance in brain alters dopamine turnover and causes behavioral disorders. Proc. Natl. Acad. Sci. 112, 3463–3468, DOI: 10.1073/pnas.1500877112 (2015).

63. de M. Lyra e Silva, N. et al. Insulin resistance as a shared pathogenic mechanism between depression and type 2 diabetes. Front. Psychiatry 10, DOI: 10.3389/fpsyt.2019.00057 (2019).

64. Bai, X., Liu, Z., Li, Z. & Yan, D. The association between insulin therapy and depression in patients with type 2 diabetes mellitus: a meta-analysis. BMJ Open 8, e020062, DOI: 10.1136/bmjopen-2017-020062 (2018).

65. Gorard, D. A., Gomborone, J. E., Libby, G. W. & Farthing, M. J. Intestinal transit in anxiety and depression. Gut 39, 551–555, DOI: 10.1136/gut.39.4.551 (1996).

66. Lurie, I., Yang, Y.-X., Haynes, K., Mamtani, R. & Boursi, B. Antibiotic exposure and the risk for depression, anxiety, or psychosis. The J. Clin. Psychiatry 76, 1522–1528, DOI: 10.4088/jcp.15m09961 (2015).

