## Supplementary Files - Description for "Cross-continental environmental and genome-wide association study on children and adolescent anxiety and depression"

**ABCD Variables of Interest**

1) Air Pollution(N=11178) (Higher- Bad, Lower- Good)

• 3 years average of ground level NO2.

• Annual average of PM 2.5 at primary residential address.

2) Population Density(N=11181) (Higher- Bad, Lower- Good)

• Unadjusted population density

• Gross residential density

3) Area Crime (N = 11181) (Higher- Bad, Lower- Good)

• Uniform Crime Reports: total adult offenses

• Uniform Crime Reports: drug sale total

• Uniform Crime Reports: drug possession total

• Uniform Crime Reports: drive under influence

4) Neighborhood Safety(N=11831) (Higher- Good, Lower- Bad)

• I feel safe walking in my neighborhood, day or night.

• Violence is not a problem in my neighborhood.

• My neighborhood is safe from crime.

5) School Risk(N=11848) (Higher- Good, Lower- Bad)

• Students have lots of chances to help decide things like class activities and rules.

• I get along with my teachers.

• I feel safe at my school.

• My teacher(s) notices when I am doing a good job and lets me know.

6) Household Income(N=10851)

• Income [0, 1= Less than $5,000; 2=$5,000 through $11,999; 3=$12,000 through $15,999; 4=$16,000 through $24,999; 5=$25,000 through $34,999; 6=$35,000 through $49,999; 7=$50,000 through $74,999; 8=$75,000 through $99,999; 9=$100,000 through $199,999; 10=$200,000 and greater]

7) Family Conflict(N=11849) (Higher- Bad, Lower- Good) – Some variables are reversed to get the sum

• We fight a lot in our family.

• Family members rarely become openly angry.

• Family members sometimes get so angry they throw

things.

• Family members hardly ever lose their tempers.

• Family members often criticize each other.

• Family members sometimes hit each other.

• If there’s a disagreement in our family, we try hard

to smooth things over and keep the peace.

• Family members often try to one-up or outdo each

other.

• In our family, we believe you do not ever get

anywhere by raising your voice.

8) Early Life Stress(N=10505) (Higher- Bad, Lower- Good) – Some variables were reversed to get the sum

In case of Early Life Stress (ELS), six categories were taken in consideration: Physical abuse/Expose or experience to trauma/Sexual Abuse; Household substance abuse; Household mental illness; Criminal in household; Parent separation /divorce; Emotional Neglect.

**cVEDA Variables of Interest**

1) Air Pollution:

- Air conditioning : Air Pollution
- Ventilation of the dwelling unit : Air Pollution

2) Area Education:

- SDI_07= Education [years]
- SDI_08 = Educational level :
- SDI_09= Did the subject never enroll/discontinue/drop out of school/college?

3) Family Conflicts:

- Sum of (IFVCS_CONTROL_1,IFVCS_CONTROL_10,IFVCS_CONTROL_11,IFVCS_CONTROL_12,IFVCS_CONTROL_13,IFVCS_CONTROL_14,IFVCS_CONTROL_2,IFVCS_CONTROL_3,IFVCS_CONTROL_4,IFVCS_CONTROL_5,IFVCS_CONTROL_6,IFVCS_CONTROL_7,IFVCS_CONTROL_8,IFVCS_CONTROL_9) **ControlScore**
- Sum of (IFVCS_PHYSICAL_1,IFVCS_PHYSICAL_10,IFVCS_PHYSICAL_11,IFVCS_PHYSICAL_12,IFVCS_PHYSICAL_13,IFVCS_PHYSICAL_14,IFVCS_PHYSICAL_15,IFVCS_PHYSICAL_16,IFVCS_PHYSICAL_2,IFVCS_PHYSICAL_3,IFVCS_PHYSICAL_4,IFVCS_PHYSICAL_5,IFVCS_PHYSICAL_6,IFVCS_PHYSICAL_7,IFVCS_PHYSICAL_8,IFVCS_PHYSICAL_9) **PhysicalAbuse**
- Sum of (IFVCS_PSYCH_1,IFVCS_PSYCH_10,IFVCS_PSYCH_11,IFVCS_PSYCH_12,IFVCS_PSYCH_13,IFVCS_PSYCH_14,IFVCS_PSYCH_15,IFVCS_PSYCH_16,IFVCS_PSYCH_17,IFVCS_PSYCH_18,IFVCS_PSYCH_19,IFVCS_PSYCH_2,IFVCS_PSYCH_20,IFVCS_PSYCH_21,IFVCS_PSYCH_22,IFVCS_PSYCH_3,IFVCS_PSYCH_4,IFVCS_PSYCH_5,IFVCS_PSYCH_6,IFVCS_PSYCH_7,IFVCS_PSYCH_8,IFVCS_PSYCH_9) **PsychologicalAbuse**
- Sum of (IFVCS_SEXUAL_1,IFVCS_SEXUAL_10,IFVCS_SEXUAL_11,IFVCS_SEXUAL_2,IFVCS_SEXUAL_3,IFVCS_SEXUAL_4,IFVCS_SEXUAL_5,IFVCS_SEXUAL_6,IFVCS_SEXUAL_7,IFVCS_SEXUAL_8,IFVCS_SEXUAL_9) **SexualAbuse** have

4) School Risk:

- Sum of (SCQ_01,SCQ_05R,SCQ_09R,SCQ_13R,SCQ_17R) SCQ_SAFETY_ORDER
- Sum of (SCQ_02,SCQ_06,SCQ_10,SCQ_14,SCQ_18) SCQ_SUPPORT_ACCEPTANCE
- Sum of (SCQ_03,SCQ_07,SCQ_11,SCQ_15,SCQ_19) SCQ_EQUITY_FAIRNESS
- Sum of (SCQ_04,SCQ_08,SCQ_12,SCQ_16,SCQ_20,SCQ_21) SCQ_ENCOURAGING_AUTONOMY haven_labelled

5) Early Life Stress
i. Physical abuse
   - Did a parent, guardian, or other household member yell, scream or swear at you, insult or humiliate you?
   - Did a parent, guardian or other household member spank, slap, kick, punch or beat you up?
   - Did a parent, guardian or other household member hit or cut you with an object, such as a stick (or cane), bottle, club, knife, whip etc.?
   - Has anything really awful happened to you? Like being in a flood, tornado, or earthquake?
     - Like being in a fire or a really bad accident? Like seeing someone get killed or hurt really bad. Like being attacked by someone?
ii. Sex abuse
    - Did someone touch or fondle you in a sexual way when you did not want them to?
    - Did someone make you touch their body in a sexual way when you did not want them to?
    - Did someone attempt oral, anal, or vaginal intercourse with you when you did not want them to?
    - Did someone actually have oral, anal, or vaginal intercourse with you when you did not want them to?
iii. Household Substance abuse

- Were your parents/guardians too drunk or intoxicated by drugs to take care of you?

iv. Household mental illness

- Did you live with a household member who was depressed, mentally ill or suicidal?

v. Parent separation/divorce

- Were your parents ever separated or divorced?
- Did your mother, father or guardian die?

vi. Family member been to jail

- Did you live with a household member who was ever sent to jail or prison? : FAMILY

vii. Expose or experience to trauma

- Did you see or hear someone being stabbed or shot in real life?
- In the past month did you:Have any accident?

viii. Emotion neglect

- Did a parent, guardian or other household member threaten to, or actually, abandon you or throw you out of the house?
- Did you feel that there was someone to take care of you and protect you?
- Did your family make you feel important or special?
- Did your parents/guardians understand your problems and worries?

**IMAGEN Variables of Interest**

1) Early Life Stress Questions
   - Parents divorced
   - Family accident or illness
   - Death in family
   - Face broke out with pimples
   - Brother or sister moved out
   - Started seeing a therapist
   - Parent changed jobs
   - Got or made pregnant
   - Thought about suicide
   - Got or gave sexually transmitted disease
   - Family had money problems
   - Parents argued or fought
   - Broke up with boy/ girlfriend
   - Family moved
   - Serious accident or illness
   - Parent abused alcohol
   - Fire (Parent1)
   - Other disasters (Parent1)
   - Attack or threat (Parent1)
   - Physical abuse (Parent1)
   - Sexual abuse (Parent1)
   - Rape (Parent1)
   - Witnessed domestic violence (Parent1)
   - Witnessed attack (Parent1)
   - Witnessed accident, sudden death (Parent1)
   - PTSD: Other severe trauma (Parent1)
2) School Risk

- I was bullied at school (a student/ peer said or did nasty or unpleasant things to me).
- I was called mean names, was made fun of, or teased in a hurtful way by a student/ peer.
- A student/ peer left me out of things on purpose, excluded me from their group of friends or completely ignored me.
- I was hit, kicked, pushed or shoved around, or locked indoors by a student/ peer.
- I took part in bullying another student/ peer at school.
- I called another student/ peer mean names, made fun of, or teased him or her in a hurtful way.'
- I kept a student/ peer out of things on purpose, excluded that student/peer from my group of friends, or completely ignored that student/ peer.
- I hit, kicked, pushed, shoved around, or locked a student/ peer indoors.
- I have been bullied by a teacher.
- I have bullied a teacher.

3) Family Conflicts

*MEAN(leq_01_feel,leq_22_feel,leq_24_feel,leq_34_feel,leq_39_feel)'.*

variable labels p1fs10 'Family stresses: Tension with partner (Parent1)'.

**Data Harmonization Figures**

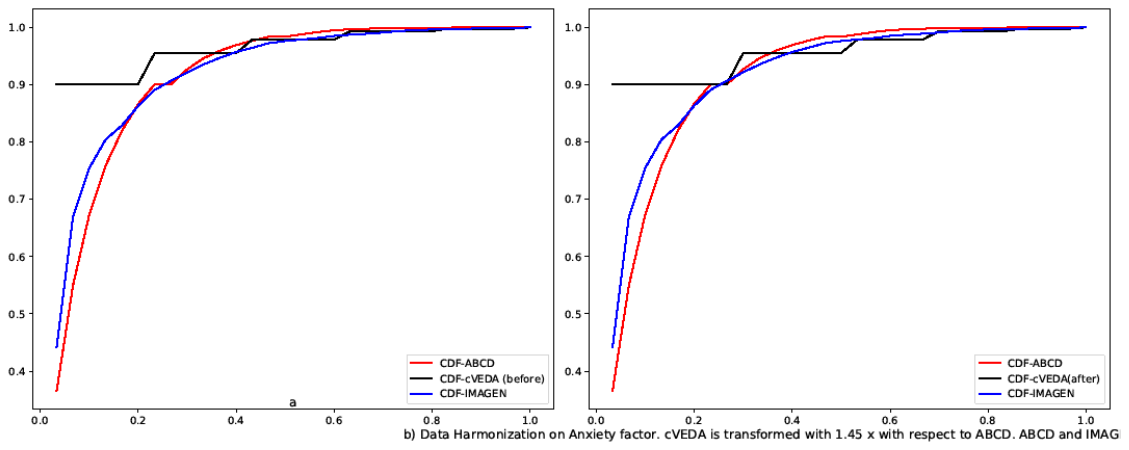

Supplementary Figure 1: Data Harmonization of Anxiety factor for cVEDA and IMAGEN with respect to ABCD. cVEDA is transformed with 1.45x

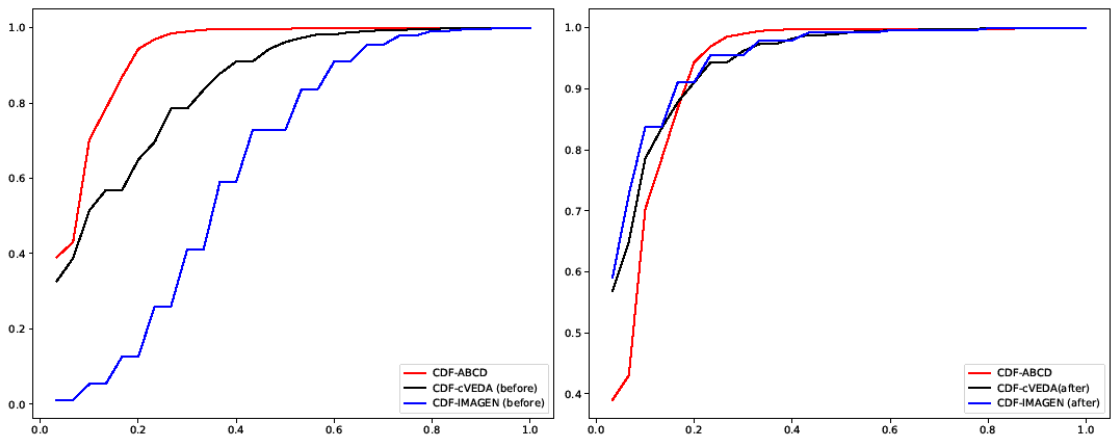

Supplementary Figure 2: Data Harmonization of ELS factor for cVEDA and IMAGEN with respect to ABCD. cVEDA is transformed with 1.75x, and IMAGEN is transformed with 3.5x.

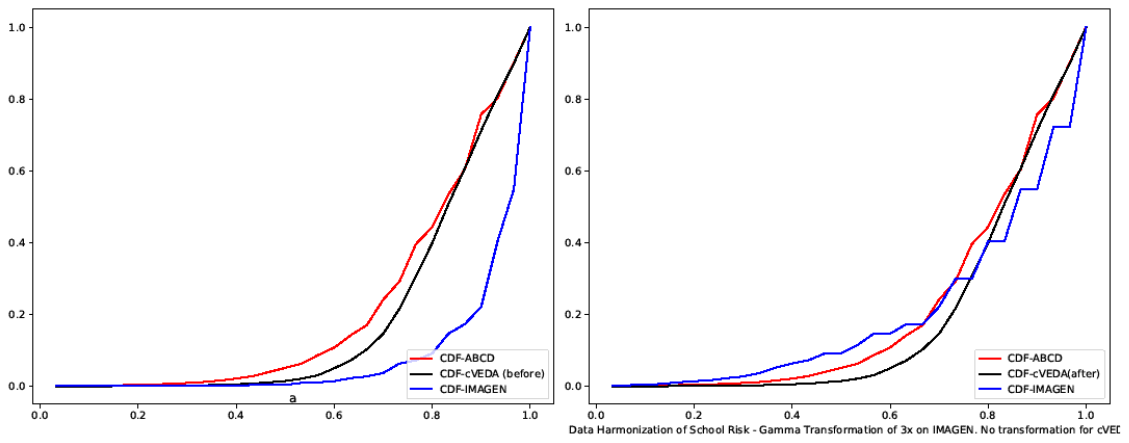

Supplementary Figure 3: Data Harmonization of School factor for cVEDA and ABCD with respect to ABCD. IMAGEN is transformed with 3x. No transformation for cVEDA.

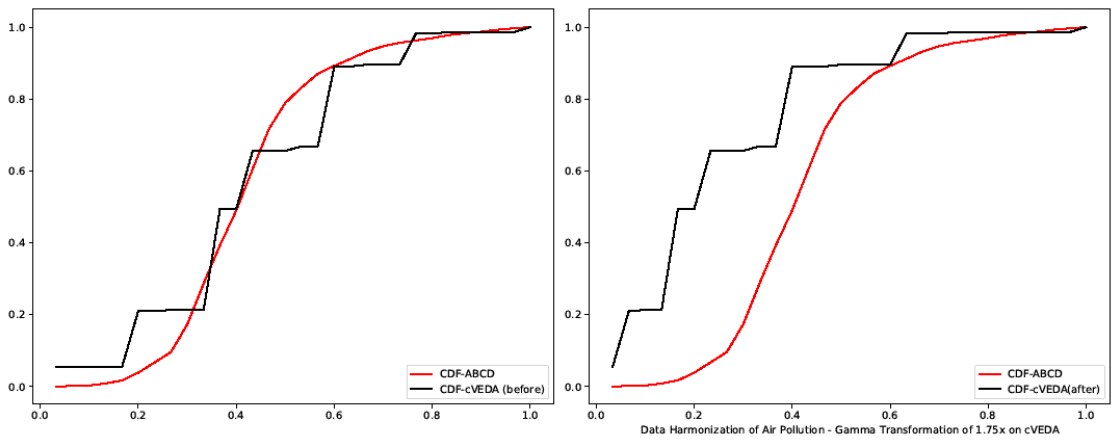

Supplementary Figure 4: Data Harmonization of Air Pollution for cVEDA with respect to ABCD. cVEDA is transformed with 1.75x.

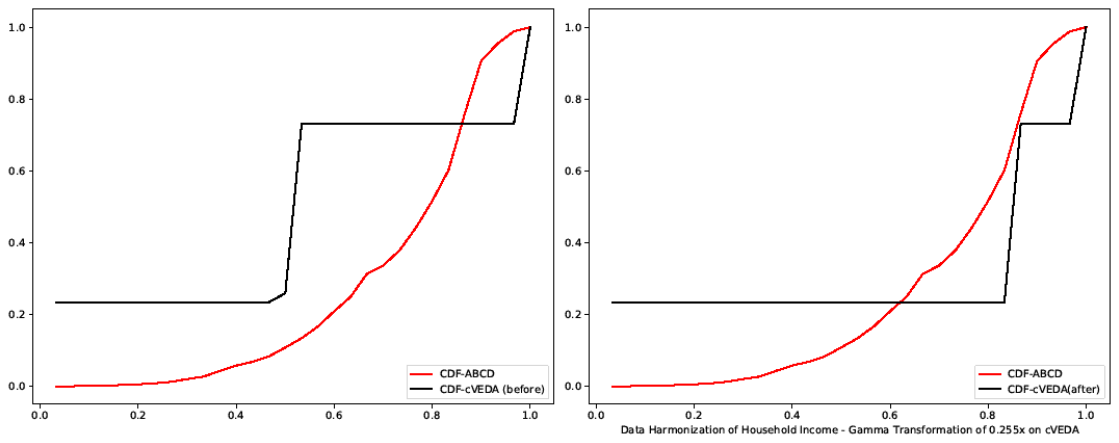

Supplementary Figure 5: Data Harmonization of Household Income for cVEDA with respect to ABCD. cVEDA is transformed with 0.255x.
